# Health Information Exchanges (HIEs) as Novel Sources for Population-Based Post Marketing Surveillance of Medical Products: A Pilot Study from the FDA Sentinel Innovation Center

**DOI:** 10.1101/2025.08.13.25333515

**Authors:** Anjum Khurshid, David Kendrick, Haritha S. Pillai, Jenny Sun, Eliel Oliveira, Alexis Jaquish, Sarah K. Dutcher, Patricia Bright, Youjin Wang, Jie Li, Tiffany Austin, Patrick J. Heagerty, Michael E. Matheny, Sebastian Schneeweiss, Rishi J. Desai

**Affiliations:** Department of Population Medicine, Harvard Pilgrim Health Care Institute and Harvard Medical School, Boston, MA; MyHealth Access Network, Tulsa, OK, USA; Division of Pharmacoepidemiology and Pharmacoeconomics, Department of Medicine, Brigham and Women’s Hospital and Harvard Medical School, Boston, MA, USA; Connxus, Austin, TX, USA; Office of Surveillance and Epidemiology, Center for Drug Evaluation and Research, United States Food and Drug Administration, Silver Spring, MD, USA; Department of Biostatistics, University of Washington, Seattle, WA, USA; Department of Biomedical Informatics, Vanderbilt University Medical Center, Nashville, TN, USA

## Abstract

**Introduction:** Health information exchanges (HIEs) provide the capability to electronically move health care information among different health care information systems and store these data for downstream use cases. However, use of data from HIEs for postmarketing surveillance of medical products is not previously explored.

**Objectives:** To conduct a pilot descriptive study characterizing data from MyHealth Access Network - a statewide HIE for Oklahoma, to understand its utility for conducting pharmacoepidemiology studies.

**Materials and methods:** MyHealth Access Network connects 95% of all hospital activity, 100% of federally qualified health center activity, and most community behavioral health clinics, tribal health systems, and independent providers in the state. As a use case to understand data in MyHealth Access Network, we characterized patients with Type 2 Diabetes Mellitus (T2DM), aged 18 years or older and treated with one of two common antidiabetic drug classes: Sodium-Glucose Co-Transporter 2 inhibitors (SGLT-2i) and Dipeptidyl Peptidase 4 inhibitors (DPP-4i) in a new user cohort design. A cohort entry date was defined based on records of medication initiation for either of the two drug classes, using dispensing data from insurance claims (when available) or prescribing data from electronic health records (EHRs) when claims were not available. Patient characteristics including demographics, comorbidities, comedications, clinical and lifestyle related factors, and healthcare utilization factors were summarized for the two exposure groups.

**Results:** A total of 76,018 DPP-4i initiators and 101,599 SGLT-2i initiators met our inclusion criteria. The mean age was 59 years and nearly half were women in both the groups. While there was a large proportion of those with missing race information (32% DPP-4i, 24% SGLT-2i), a majority were White (47% DPP-4i, 57% SGLT-2i). The second most reported race category was American Indian or Alaskan Natives (13% DPP-4i, 9.9% SGLT-2i), which is in line with their representation in the 2024 US Census for Oklahoma. Hemoglobin A1c (HbA1c) results were available for 63% and 66% of the DPP-4i and SGLT-2i groups with mean + SD of 8.1+ 1.9% and 8.2 + 1.9%, respectively. Serum creatinine was recorded for 67% DPP-4i and 70% SGLT-2i initiators with mean + SD of 1.30 + 1.01 mg/dL and 1.14 + 0.64 mg/dL, respectively.

**Discussion:** HIEs may offer a promising resource for population-based postmarketing surveillance of medical products owing to their comprehensive capture of information across various healthcare settings.

## 1. Introduction

Fragmentation in the U.S. healthcare system poses critical challenges for both patient level care coordination and population level research. A 2007 analysis of Medicare data reported that patients visited, on average, more than 7 different healthcare providers annually, and that, on average, primary care providers serving a panel of patients found themselves coordinating care among 225 other providers in 117 other organizations.[1] As the adoption of electronic health records (EHR) by eligible providers and hospitals in the U.S. healthcare system surged following significant investments through the Health Information Technology for Economic and Clinical Health (HITECH) Act, the need to facilitate seamless exchange of clinical data securely across disparate competing EHR systems and provider organizations was clearly recognized.[2] As described in the Office of the National Coordinator for Health IT (ONC) Roadmap for Nationwide Interoperability (2015)[3], “every patient deserves to have their complete, longitudinal health record available where and when they need it for decisions about their care”. As a result, approximately 3% of HITECH funding supported the dramatic expansion of state and regional health information exchanges (HIEs) throughout the country.[4] The primary purposes of HIEs is to promote interoperability and coordination of care. Implementing and operationalizing this mission has been done by creating trusted regional networks where the silos of data, existing due to the fragmented nature of healthcare delivery, are connected through a governed, secure, central exchange platform. Since many of these exchanges also maintain the exchanged clinical documents and patient records in a centralized clinical data repository, over the years, these HIEs have become a comprehensive source of longitudinal, population level, linked EHR data in the country.[5]

Almost all states in the U.S. have a statewide HIE or multiple regional HIEs. A recent survey funded by the ONC has reported about 80 fully operational HIEs in the country with an average data repository comprising more than four million unique patients with data going back to around 15 years in some cases. [6,7] These HIEs are mostly non-profit entities with a multistakeholder community governance. They maintain a robust technical infrastructure to create bidirectional data interfaces with large and small hospitals, federally qualified health clinics, physician practices, Emergency Medical Services (EMS), behavioral health providers, pharmacies, laboratories, and public health clinics among other types of organizations that are sources of health-related data. These interfaces allow for exchange of detailed clinical data through secure connections using FHIR APIs (Fast Health Interoperability Resources Application Programming Interfaces), HL7 (Health Level 7) interfaces, CCD (Continuity of Care Documents), or even flat files (.csv).[8] Importantly, HIEs maintain governance and compliance required to build trust in the privacy and confidentiality of sensitive clinical data. The HIEs store and use this data as Business Associate Agreement (BAA) partners for all providers connected to them and fully comply with HIPAA (Health Insurance Portability and Accountability Act, 1996) and other federal, state and local laws.[9] There are also continuous data quality and data integrity assurance processes[10] implemented in all major HIEs. This broad technical infrastructure has matured over the last decade, making HIEs an attractive source of large-scale EHR data that is ready to be leveraged for multiple important use cases. However, historically HIEs have been underutilized for conducting public health surveillance, pharmacoepidemiologic studies, and population outcomes research.[11]

The Sentinel Initiative is a key component of the U.S. Food and Drug Administration’s (FDA) postmarket surveillance for approved medical products. Results from Sentinel investigations are routinely used to support the FDA in regulatory decision making.[12] The Sentinel System, a national medical product safety surveillance system within the Sentinel Initiative, has historically relied on large volumes of insurance claims data but more recently has developed an infrastructure linking EHRs with insurance claims to address questions related to medical product use and associated outcomes using more granular clinical information captured in EHRs.[13,14] The new infrastructure, the Real World Evidence Data Enterprise (RWE-DE), comprises linked claims-EHR data for 26 million individuals from six data sources involving four academic partner institutions (three tertiary care centers-Mass General Brigham, Vanderbilt University Medical Center, Duke University Health Systems, and an integrated care delivery network-Kaiser Permanente-Washington) collectively termed the Development Network and two commercial data networks (HealthVerity, TriNetX) collectively termed the Commercial Network.[14] While a major step forward, the RWE-DE may benefit from inclusion of additional data sources to increase the total population covered and availability of near-real time information. Additionally, all current sources face challenges in EHR data continuity due to the fragmentation of care described above. To explore the potential of HIEs for addressing some of these challenges, we initiated a pilot study with MyHealth Access Network-a statewide HIE from Oklahoma, to implement a descriptive analysis of initiators of two antidiabetic medications to reflect a typical Sentinel query. In this article, we describe the learnings from the pilot and discuss considerations for future use of HIEs as novel sources for population-based pharmacoepidemiology research and postmarketing surveillance of medical products.

## 2. Methods

### 2.1 Data source

MyHealth Access Network is a non-profit organization and Oklahoma’s State Designated Entity for Health Information Exchange that connects 95% of all hospital activity, 100% of federally qualified health center activity, and most community behavioral health clinics, tribal health systems, and independent providers in the state.[15] It connects to more than 500 unique healthcare entities including independent pharmacies, emergency medical services (EMS), social services organizations, organ procurement organizations, Oklahoma Office of the Chief Medical Examiner, and health plans in the area. The HIE has a clinical data repository that goes back to encounters captured since 2011 and provides the most complete and near real-time access to population-level healthcare and social needs data for the over 4 million residents of Oklahoma. These data are standardized and linked across all providers into a unique unduplicated record for each patient. We used data from January 2016 to December 2023 available in MyHealth Access Network, which also included linked EHR data to claims data from three of the health plans.

### 2.2 Study design

We conducted a descriptive medical product utilization query (similar to a ‘Type 5’ query in the Sentinel Routine Querying tools)[16]-to identify and characterize patients with Type 2 Diabetes Mellitus (T2DM) treated with two common antidiabetic drug classes: Sodium-Glucose Co-Transporter 2 inhibitors (SGLT-2i) and Dipeptidyl Peptidase 4 inhibitors (DPP-4i) in a new user cohort design. Patients entered the cohort on a cohort entry date (CED) based on records of medication initiation for either of the two drug classes, using dispensing data from insurance claims (when available) or prescribing data from EHRs when claims were not available. Patients were required to be aged 18 years or older, have a diagnosis of type 2 diabetes mellitus, and have some EHR activity (operationalized as having any data available from EHRs any time prior to index date) to ensure adequate contact with the healthcare system and capture of clinical codes to measure baseline characteristics. Since the focus of this query was on patients with type 2 diabetes mellitus, we excluded patients with a diagnosis of type 1 diabetes mellitus.

### 2.3. Variables

We assessed demographic characteristics (age, sex, race, ethnicity and year of cohort entry); diabetes-related factors including duration of diabetes (operationalized as the time between the first diagnosis codes found in the data and cohort entry) and use of other antihyperglycemic medications; a range of comedications and comorbid conditions; laboratory test results and other clinical parameters including serum creatinine, hemoglobin A1c (HbA1c), total cholesterol, high density lipoprotein (HDL), low density lipoprotein (LDL), and left ventricular ejection fraction (LVEF); vital signs (heart rate, blood pressure), body mass index (BMI), and lifestyle factors (any smoking status, alcohol use, drug misuse or abuse). All variables were assessed in a 365-day baseline period prior to the cohort entry date, unless otherwise specified. For laboratory tests, clinical parameters, and vital signs, values closest to the cohort entry were used for patients with multiple recordings in the baseline period. Standardized coding terminology including national drug codes (NDCs) and RxNorm Concept Unique Identifier (RxCUI) for medications with drug mechanism classifications determined by manual mapping, Logical Observational Identifiers Names and Codes (LOINC) for laboratory tests, international classification of diseases (ICD)-10-CM codes for diagnoses, and current procedural terminology (CPT) codes (with modifiers) to assess in-person healthcare utilization were used to identify relevant information in each domain.

### 2.4 Statistical analysis

We used basic descriptive statistics to summarize all characteristics for each of the two exposure groups using percentages for categorical variables and means (standard deviations) for quantitative measures. We also reported the proportion of individuals in each group who had at least one in-person EHR encounter in 30, 90, 180, and 365 days prior to the cohort entry to contextualize routine flow of information in the HIE.

### 2.5 Ethics review

This Sentinel project is a public health surveillance activity conducted under the authority of the Food and Drug Administration and, accordingly, is not subject to Institutional Review Board oversight.[17-19] Additionally, the HIE governance committee comprising local stakeholders and data contributors, reviewed and approved the secondary use of the data for this query.

## 3. Results

Figure 1. shows the total number of DPP-4i and SGLT-2i initiators with type 2 diabetes mellitus along with source of the prescription information on their cohort entry date. A total of 76,018 DPP-4i initiators and 101,599 SGLT-2i initiators met our inclusion criteria, a majority of whom were included based on prescription orders found in EHRs. **Figure 2** outlines information sources for the two exposure groups during the study period. A large majority (63.0% and 60.1% for DPP-4i and SGLT-2i groups, respectively) were identified from EHR data alone and did not appear in any of the three claims datasets. A total of 15.7% and 20.1% for DPP-4i and SGLT-2i, respectively had information in both EHRs and claims from one or more of the three payers. Notably, a small subset of patients (0.003% and 0.01% for DPP-4i and SGLT-2i) existed in all three payer datasets and EHR at some point of time for both DPP-4i and SGLT-2i initiators, which highlights the issue of frequent changes in individuals’ insurance coverage over time with claims data sources.

**Figure 1.**
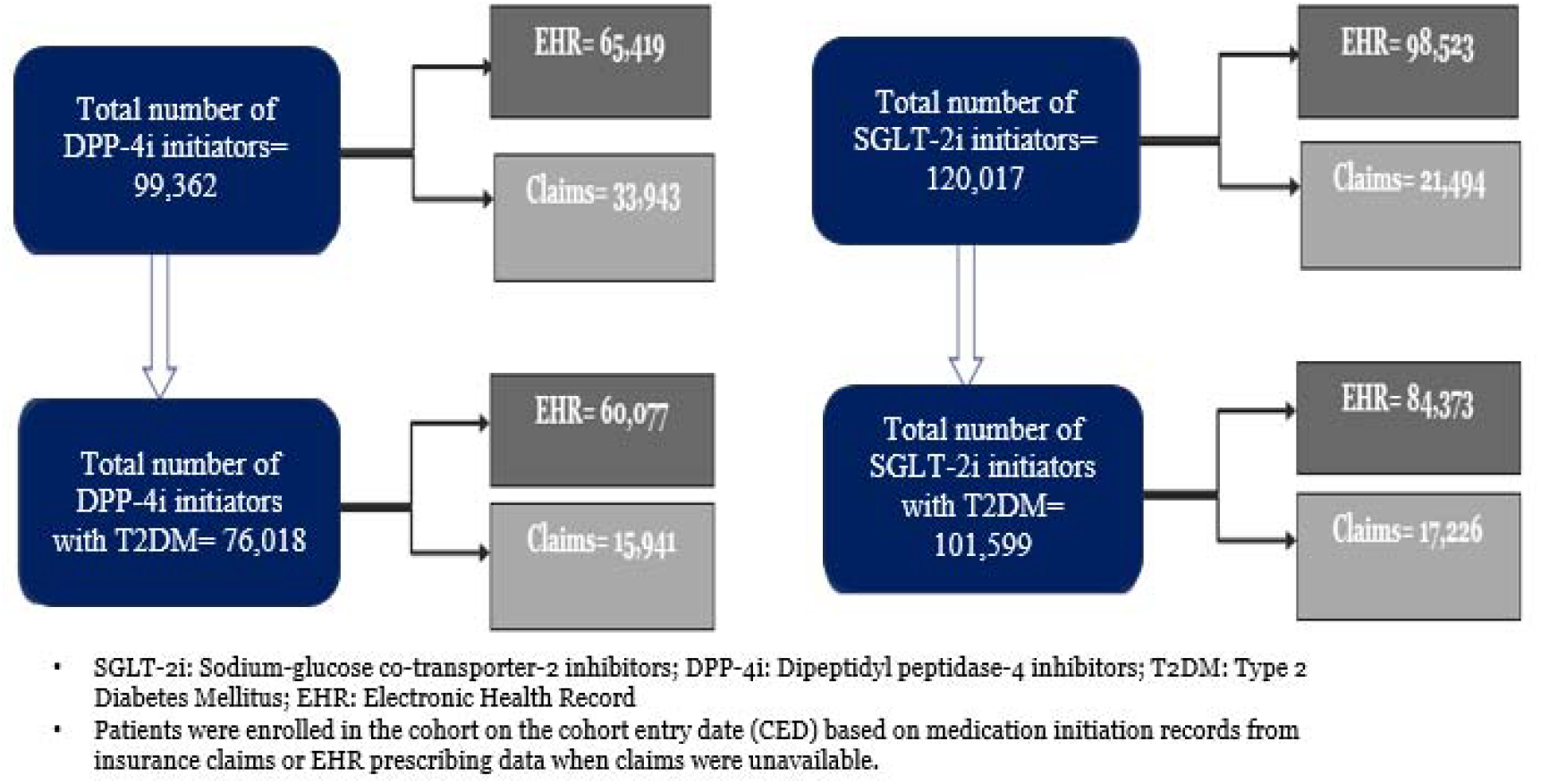
Final cohort of DPP-4i and SGLT-2i initiators with type 2 diabetes mellitus along with data source (EHR versus claims)

**Figure 2.**
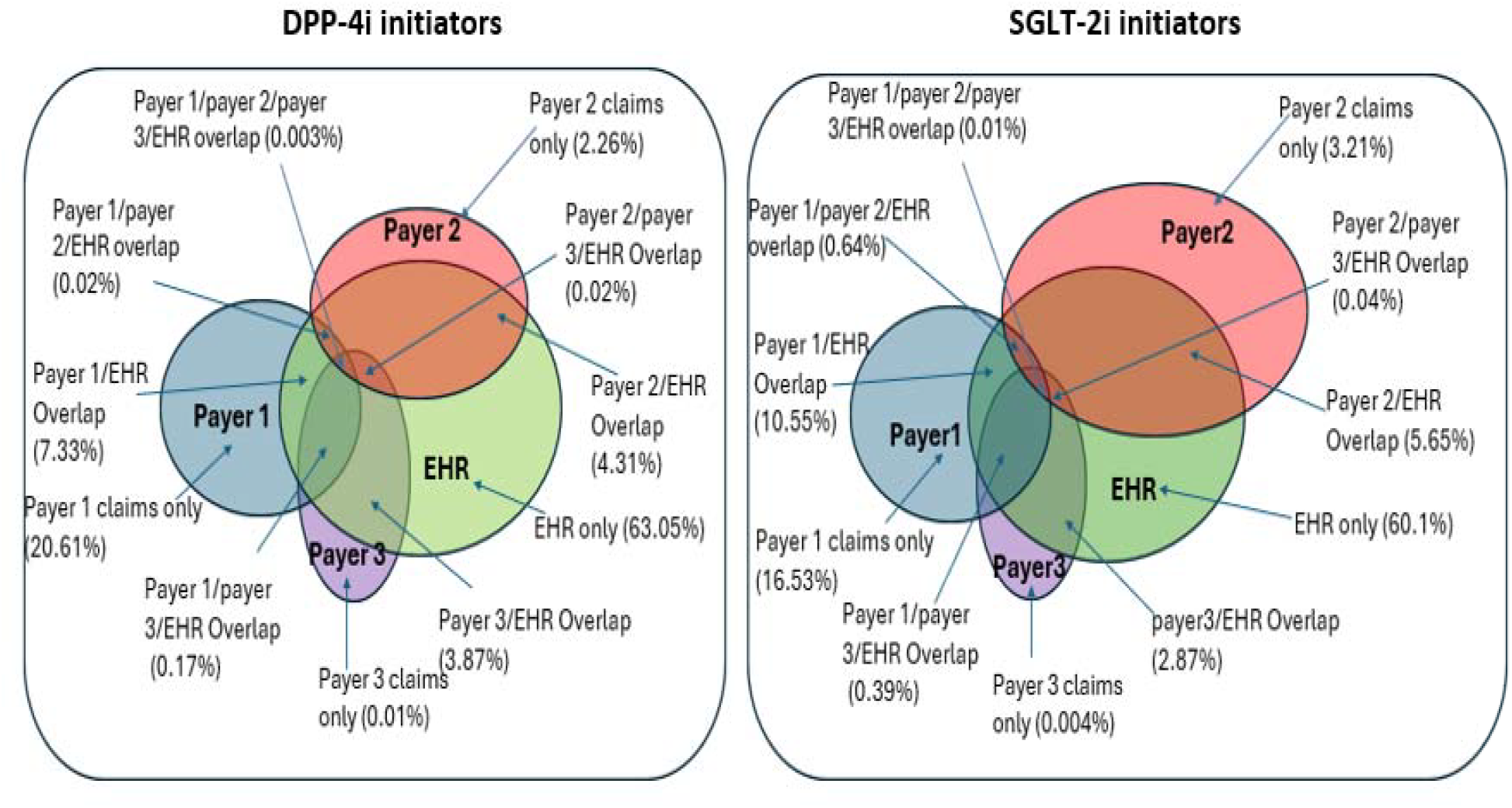
Claims versus EHR overlap of DPP-4i and SGLT-2i initiators with type 2 diabetes mellitus

### Patient demographics, medication use, and comorbid diagnoses

**Table 1** summarizes the demographic characteristics of the DPP-4i and SGLT-2i initiators. The mean age was 59 years in each of the cohorts with the largest group of patients falling within the age category of 55-64 years (30%, 32% respectively). About half of the patients were female in both the cohorts (49%, 45% respectively). Less than 0.05% of patients had missing data for age and sex. While information on race was unknown or missing for 32% of the DPP-4i initiators and 24% of SGLT-2i initiators, a majority of patients were White in both the cohorts (47%, 57% respectively); the second most reported race category was American Indian or Alaskan Natives, representing 13% and 9.9%, respectively. A total of 8.5% of the DPP-4i initiators and 7.5% of the SGLT-2i initiators were of Hispanic origin.

**Table 1.**
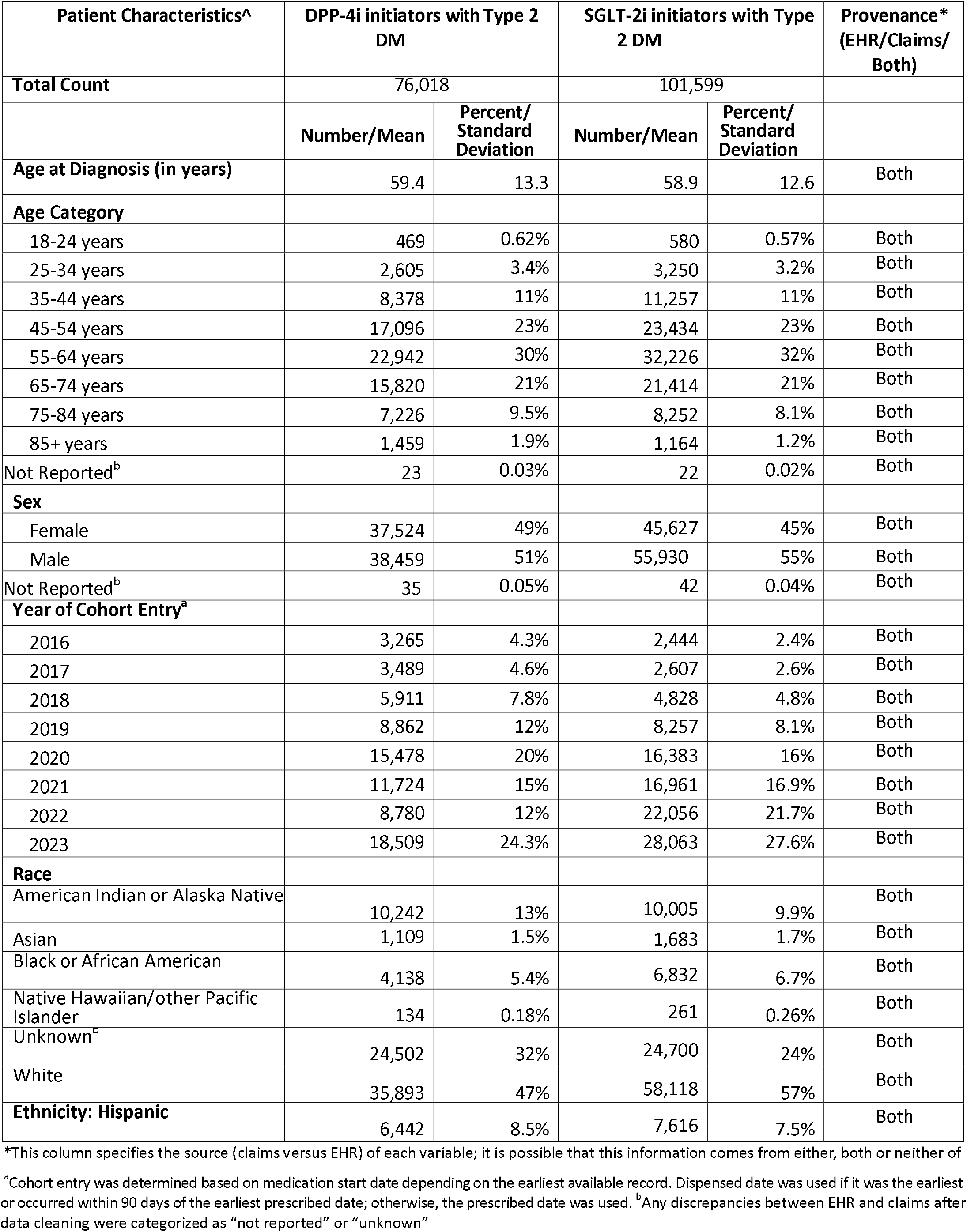
Demographic characteristics of the DPP-4i and SGLT-2i initiators with type 2 diabetes mellitus

**Table 2** provides information on diabetes duration, comedications, and comorbid conditions. Most patients had a diagnosis of Type 2 DM for 1-5 years at the time of cohort entry across both DPP-4i initiators and SGLT-2i initiators (39%, 48%). Among other antidiabetic medications, metformin was most common, followed by sulfonylureas. GLP1 agonists were much more frequently used by SGLT-2i initiators (19% vs 6.3%). Angiotensin converting enzyme inhibitors and angiotensin receptor blockers were the most common co-medications in both the groups (42%, 52%) and hypertension was the most frequent comorbid diagnosis (32%, 45%).

**Table 2.** Diabetes duration, comedications, and comorbidities observed in the DPP-4i and SGLT-2i initiators with type 2 diabetes mellitus

| Patient Characteristics | DPP-4 inhibitor initiators with Type 2 DM |  | SGLT-2 inhibitor initiators with Type 2 DM |  | Provenance* (EHR/Claims/Both) |
| --- | --- | --- | --- | --- | --- |
|  | N/Mean | %/SD | N/Mean | %/SD |  |
| **Duration of diabetes (in years), N (%)<sup>a</sup>** |  |  |  |  |  |
| < 1 | 26,599 | 35% | 32,330 | 32% | Both, although EHR had longer history |
| 1-5 | 29,323 | 39% | 48385 | 48% | Both, although EHR had longer history |
| 6-10 | 4,549 | 6.0% | 9920 | 9.8% | Both, although EHR had longer history |
| 11-15 | 846 | 1.1% | 1770 | 1.7% | Both, although EHR had longer history |
| >15 | 374 | 0.49% | 398 | 0.39% | Both, although EHR had longer history |
| **<sup>b</sup>History of Antihyperglycemic Agents** |  |  |  |  |  |
| Metformin | 34,590 | 46% | 53,066 | 52% | EHR: Prescribed; Claims: Dispensed |
| Thiazolidinediones | 4,977 | 6.6% | 6,493 | 6.4% | EHR: Prescribed; Claims: Dispensed |
| Sulfonylureas | 18,679 | 25% | 23,238 | 23% | EHR: Prescribed; Claims: Dispensed |
| Alpha Glucosidase Inhibitors | 603 | 0.79% | 452 | 0.44% | EHR: Prescribed; Claims: Dispensed |
| Meglitinides | 201 | 0.26% | 255 | 0.25% | EHR: Prescribed; Claims: Dispensed |
| Glucagon-like Peptide-1 Agonists | 4,791 | 6.3% | 19,189 | 19% | EHR: Prescribed; Claims: Dispensed |
| Short/Rapid Acting Insulins | 7,023 | 9.2% | 13,247 | 13% | EHR: Prescribed; Claims: Dispensed |
| Long/Intermediate Acting Insulins | 4,100 | 5.4% | 8,624 | 8.5% | EHR: Prescribed; Claims: Dispensed |
| Long and Short Acting Insulin Combinations | 713 | 0.94% | 1,355 | 1.3% | EHR: Prescribed; Claims: Dispensed |
| **Count of total oral antihyperglycemic agents** | 0.87 | 0.96 | 1.23 | 1.06 | EHR: Prescribed; Claims: Dispensed |
| **<sup>b</sup>History of other medications** |  |  |  |  |  |
| Antiplatelets | 20,123 | 26% | 30,937 | 30% | EHR: Prescribed; Claims: Dispensed |
| Drugs for Obstructive Airway Diseases | 19,063 | 25% | 29,566 | 29% | EHR: Prescribed; Claims: Dispensed |
| Calcium Channel Blockers | 13,794 | 18% | 21,016 | 21% | EHR: Prescribed; Claims: Dispensed |
| Diuretics | 20,425 | 27% | 33,576 | 33% | EHR: Prescribed; Claims: Dispensed |
| Antidepressants | 14,962 | 20% | 24,054 | 24% | EHR: Prescribed; Claims: Dispensed |
| Beta Blockers | 21,112 | 28% | 35,158 | 35% | EHR: Prescribed; Claims: Dispensed |
| Agents Acting on the Renin-Angiotensin System | 31,585 | 42% | 52,466 | 52% | EHR: Prescribed; Claims: Dispensed |
| **History of comorbidities** |  |  |  |  |  |
| Acute Kidney Failure/Chronic Kidney Disease | 18,160 | 24% | 36,092 | 36% | Both |
| Acute Myocardial Infarction | 939 | 1.2% | 2,321 | 2.3% | Both |
| Acute Respiratory Disease | 7,569 | 10% | 14,881 | 15% | Both |
| Anemia | 4,099 | 5.4% | 7,008 | 6.9% | Both |
| Atrial Fibrillation | 2,373 | 3.1% | 5,111 | 5.0% | Both |
| Cardiomyopathy | 1,070 | 1.4% | 3,835 | 3.8% | Both |
| Chronic Obstructive Pulmonary Disorder-Like Symptoms | 3,372 | 4.4% | 6,615 | 6.5% | Both |
| Coronary Revascularization | 1,749 | 2.3% | 4,053 | 4.00% | Both |
| Depressive Disorder | 3,989 | 5.3% | 6,952 | 6.8% | Both |
| Gastroesophageal Reflux Disease | 5,456 | 7.2% | 10,029 | 9.9% | Both |
| Heart Failure | 3,094 | 4.1% | 8,970 | 8.8% | Both |
| Hypercholesterolemia/Hyperlipidemia | 18,123 | 24% | 36,712 | 36% | Both |
| Hypertension/Hypertensive Disorder | 24,154 | 32% | 45,443 | 45% | Both |
| Malignant Neoplastic Disease | 2,121 | 2.8% | 3,590 | 3.5% | Both |
| Nicotine Dependency | 3,348 | 4.4% | 6,265 | 6.2% | EHR |
| Obesity | 9,155 | 12% | 20,377 | 20% | Both |
| Osteoarthritis | 4,831 | 6.4% | 9,486 | 9.3% | Both |
| Stroke (Ischemic/Hemorrhagic) | 1,166 | 1.5% | 1,742 | 1.7% | Both |
| Thromboembolism | 659 | 0.87% | 1,294 | 1.3% | Both |
| Transient Ischemic Attack | 385 | 0.51% | 658 | 0.65% | Both |

\*This column specifies the source (claims versus EHR) of each variable; it is possible that this information comes from either, both or neither of the two sources.
<sup>a</sup> Defined as difference between first recorded diagnosis for Type 2 DM and cohort entry date
<sup>b</sup> Medication history was determined based on the earliest available record. Dispensed date was used if it was the earliest or occurred within 90 days of the earliest prescribed date; otherwise, the prescribed date was used.

**Table 3** summarizes information on laboratory test results, lifestyle factors and vital signs. HbA1c results were available for 63% and 66% of the DPP-4i and SGLT-2i groups, respectively with mean + SD of 8.1+ 1.9% and 8.2 + 1.9%. Serum creatinine was recorded for 67% of DPP-4i and 70% of SGLT-2i initiators. The mean serum creatinine values were 1.30 + 1.0 mg/dL and 1.14 + 0.6 mg/dL for DPP-4i and SGLT-2i initiators respectively. Substance use including smoking status, alcohol use, and drug misuse/abuse was recorded for 5-23% of the patients across both DPP-4i and SGLT-2i initiators. BMI was recorded for 28.4% of the DPP-4i initiators (mean = 33.4 + 8.0 kg/m^2^) and 46% of the SGLT-2i initiators (mean = 34.1 + 8.0 kg/m^2^). Heart rate and blood pressure had similar mean values across both groups, with measurements recorded for 25-35% of DPP-4i initiators, and a higher proportion (45-50%) of SGLT-2i initiators.

**Table 3.**
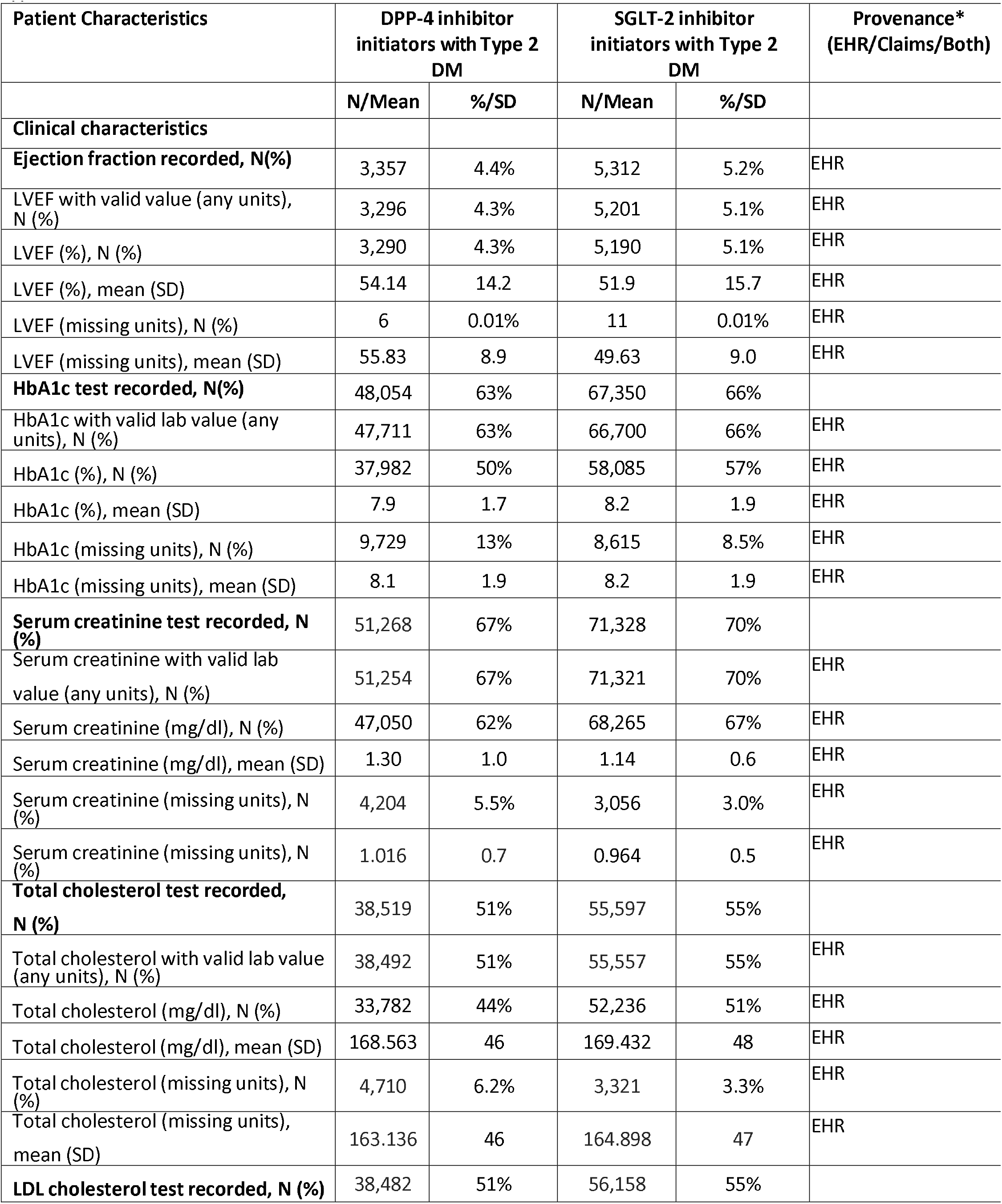
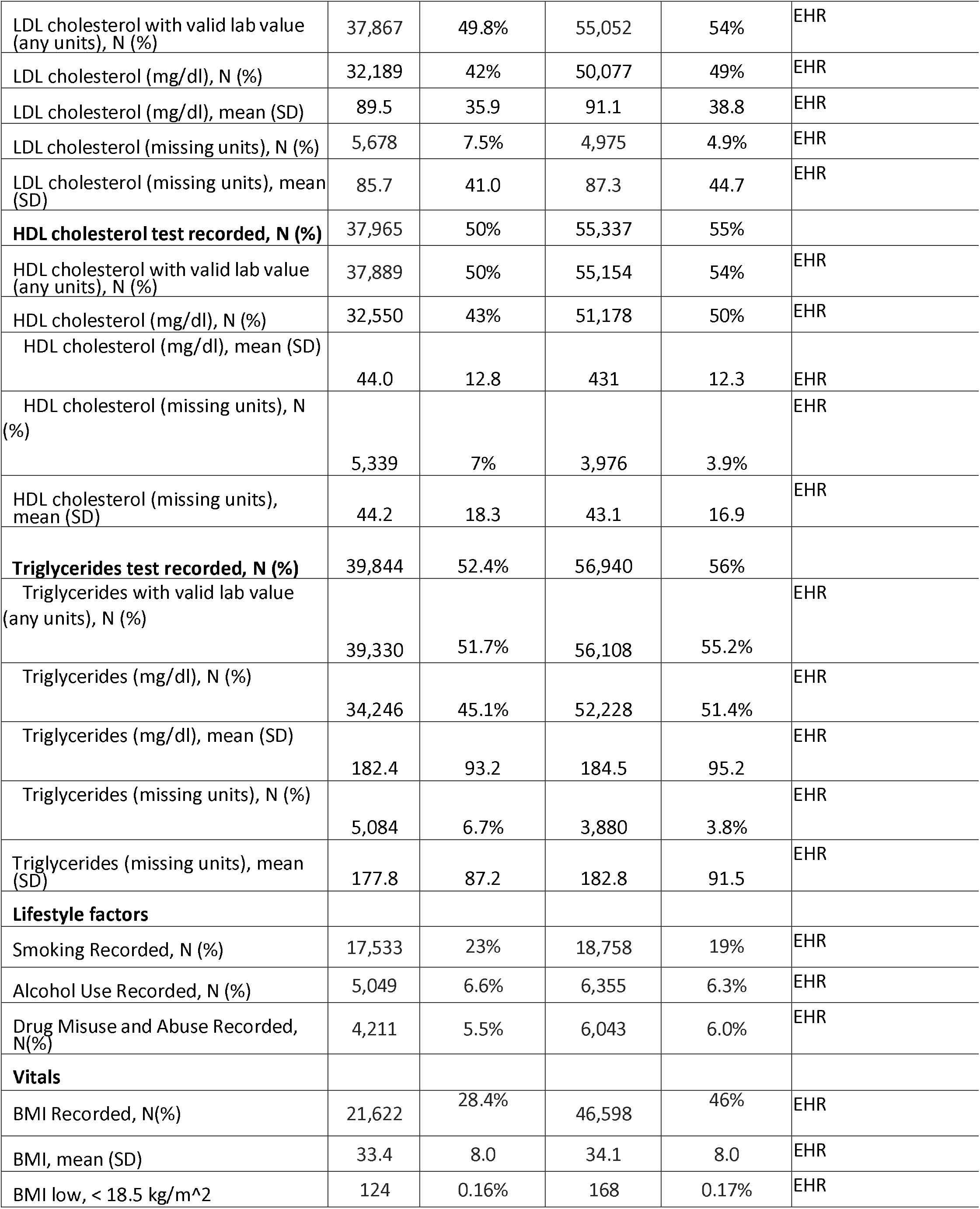
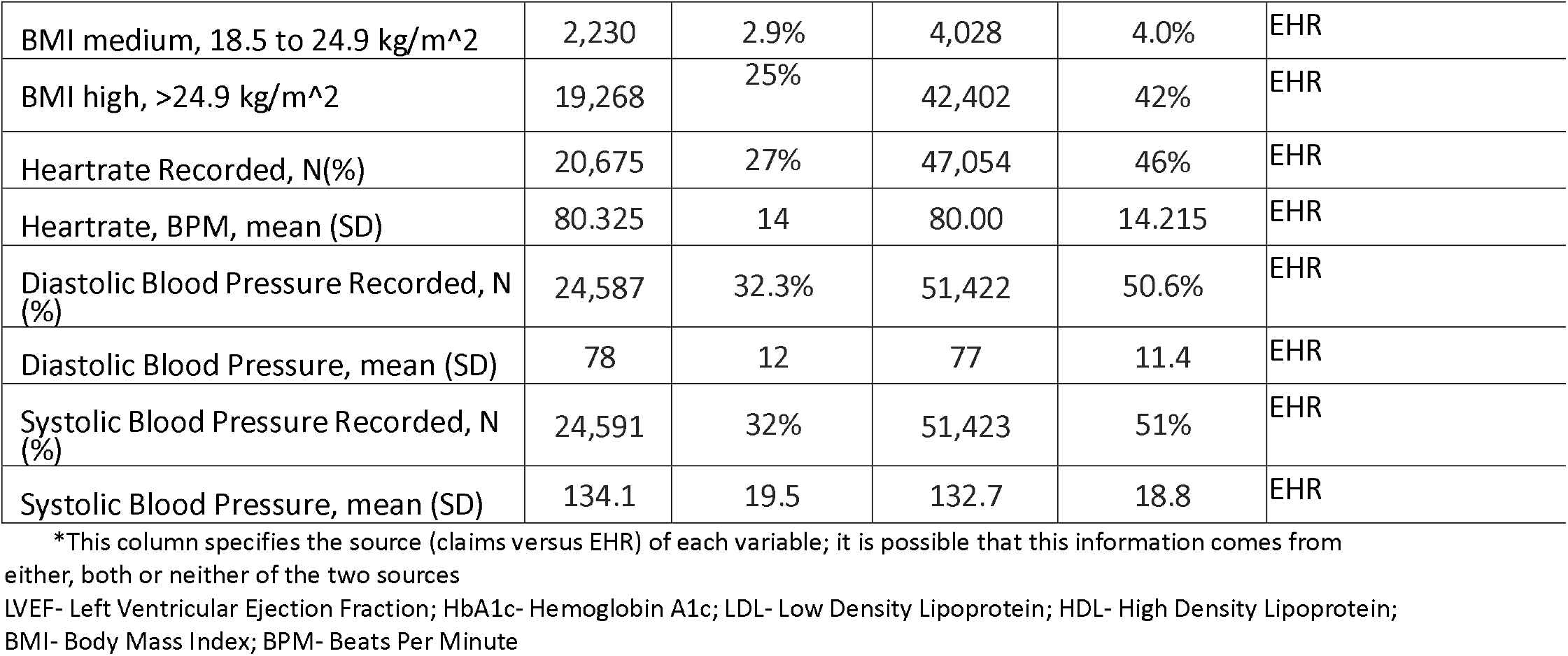
Clinical characteristics, lifestyle factors, and vital signs of the DPP-4i and SGLT-2i initiators with type 2 diabetes mellitus

**Table 4** summarizes healthcare utilization-related information. We observed that 64% of the DPP-4i initiators and 76 % of the SGLT-2i initiators had at least 1 in-person encounter in the 365 days prior to the cohort entry. This proportion was consistent when restricting to past 180 days (62 %, 74 %), 90 days (60 %, 72 %), and 30 days (55 %, 67 %).

**Table 4.**
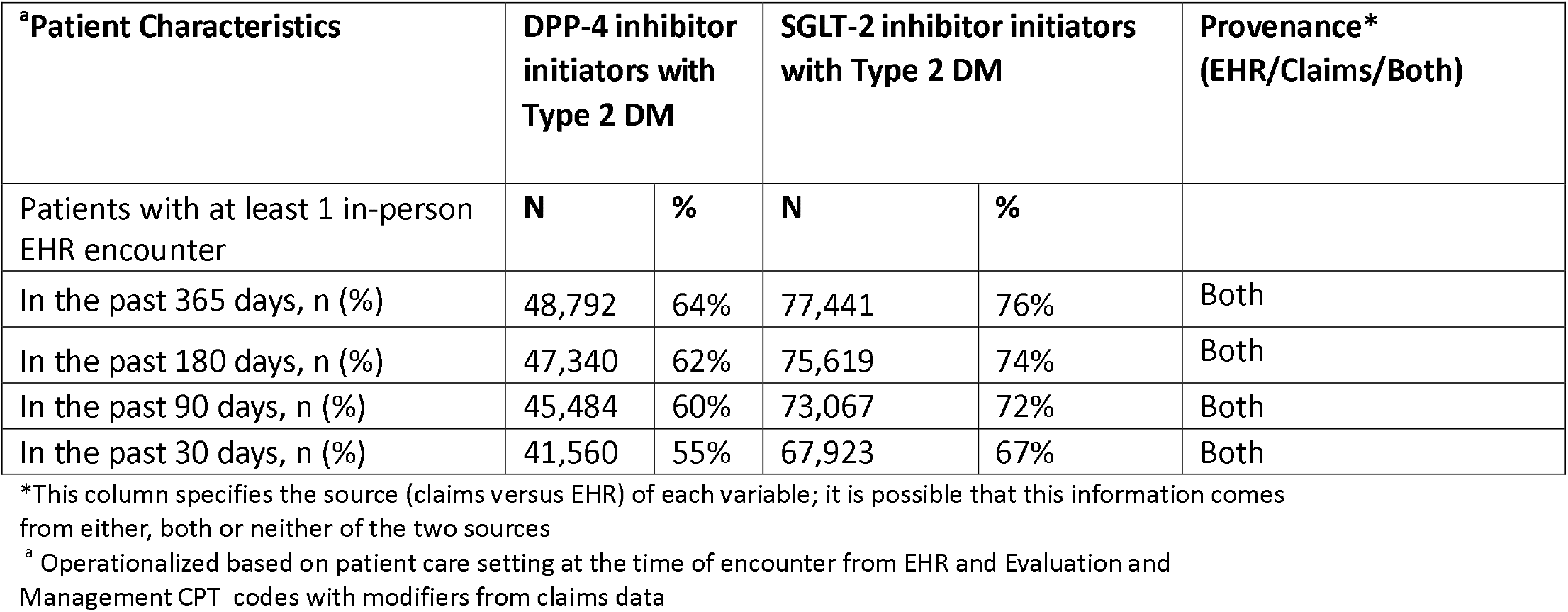
Healthcare utilization features of the DPP-4i and SGLT-2i initiators with type 2 diabetes mellitus

| <sup>a</sup>Patient Characteristics | DPP-4 inhibitor initiators with Type 2 DM |  | SGLT-2 inhibitor initiators with Type 2 DM |  | Provenance* (EHR/Claims/Both) |
| --- | --- | --- | --- | --- | --- |
| Patients with at least 1 in-person EHR encounter | N | % | N | % |  |
| In the past 365 days, n (%) | 48,792 | 64% | 77,441 | 76% | Both |
| In the past 180 days, n (%) | 47,340 | 62% | 75,619 | 74% | Both |
| In the past 90 days, n (%) | 45,484 | 60% | 73,067 | 72% | Both |
| In the past 30 days, n (%) | 41,560 | 55% | 67,923 | 67% | Both |

\*This column specifies the source (claims versus EHR) of each variable; it is possible that this information comes from either, both or neither of the two sources
<sup>a</sup> Operationalized based on patient care setting at the time of encounter from EHR and Evaluation and Management CPT codes with modifiers from claims data

## 4. Discussion

In this study, we observed that it was feasible to create patient cohorts using near real-time EHR data to describe initiators of medications using data from an HIE. We noted that capture of important clinical data elements such as HbA1c was comparable or more complete than diabetic cohorts from other EHR sources from the US.[20-22] Importantly, traditionally underrepresented racial groups such as American Indians were represented in line with their representation in the 2024 US Census for Oklahoma (10-13% in this study, 9.5% in the US Census)[23], which highlights the truly population-based nature of HIEs.

HIEs offer certain benefits that make them a promising resource for future use in national medical product surveillance systems like Sentinel for pharmacoepidemiology evaluations. First, since most HIEs are community-governed with a not-for-profit status and funding from public sources (state and federal health departments), they are quite efficient in terms of cost to acquire data for surveillance and research. As the source of almost real-time data on an entire population, HIEs may help to study emergent infections, adverse events for newly used treatments, and geographical coverage within the covered regions in a timely and comprehensive manner. Many HIEs also store unstructured data, like clinical notes, creating a valuable resource to develop and test natural language processing algorithms and large language models.[24] Increasingly, HIEs are expanding their role to collect nonmedical data such as social needs assessments, social services referrals, and criminal justice, environmental, and housing data to become health data utilities (HDUs) for their state or region, much like the energy or public transportation utilities that provide the service for multiple uses by consumers.[25] Some states have also passed legislation to mandate contribution of health and social services data to their HDUs to help state-level public health and Medicaid better coordinate, monitor, and evaluate care for its population. This rich and comprehensive data under a community-governed entity, which has high levels of technical expertise to curate representative population level data of clinical and non-clinical factors, is primed to help study health equity issues in much better detail than in any other dataset of this size and coverage.[26]

One of the most important requirements for conducting causal inference studies with longitudinal health records is the availability of EHR data without ‘information leakage’ that is common in traditional EHR sources from a single healthcare system because patients frequently seek care outside of the system.[27] A key service provided by HIEs is that of maintaining a master patient index that links all the various medical records across the fragmented health delivery system into one master record, thus saving a huge effort in deduplicating and linking clinical records for subsequent analysis. The ability to maintain a truly longitudinal patient record across healthcare systems makes HIEs an attractive data resource to conduct studies that require long term follow-up and consistent data availability over the course of a treatment or a disease. In the analysis of healthcare utilization, we observed that about 60-70% patients included in the current study had an in-person EHR visit in 365 days, 180 days, and 30 days prior to the medication initiation date, which suggests that HIEs have a stable pool of patients with consistent flow of EHR information.

Some governance related aspects of HIE data merit a discussion. While a single patient-level record can be shared across the network for treatment, payment, and operations for authorized users with an established relationship with that patient, the secondary use of these data for research and surveillance has established policies that vary across HIEs. These policies are largely focused on patient privacy and agreed upon governance policies with data contributors. Secondary use data sharing for public health, epidemiological studies, and population health research is determined on a case-by-case basis. Most HIEs will allow use of data for public health surveillance activities, but in many cases, use of data for research requires appropriate IRB supervision and may require patient consent for sharing any protected health information.

We also acknowledge some important limitations of our current work. First, we only explored a single HIE in Oklahoma which is a relatively mature and sophisticated HIE with a legislative mandate from the state. HIEs across the country differ in significant ways in terms of their geographical and clinical data capture as well as their ability to link to other sources like insurance claims. However, many of the features described for MyHealth Access are likely to be representative of larger state level and regional HIEs, some with much larger populations such as in New York, California, Michigan, and Texas. Second, we only focused on a descriptive study of relatively commonly used medication classes, which precludes generalizable conclusions regarding completeness of information for more rare conditions or medications. However, it is reasonable to hypothesize that if there is a method of connecting a consortium of HIEs from across the country to act as a distributed network with a common data model and quality assurance, as established in the Sentinel Distributed Network, this limitation may be adequately addressed through a geographically and socioeconomically representative cohort from across the country. A preliminary feasibility study for such a consortium of HIEs to support pharmacoepidemiology and population health studies is already underway and showing encouraging results.[28]

In conclusion, the HIE network has grown over the last decade at the local, regional, and national level. Our pilot study conducted in partnership with MyHealth Access Network suggests that data from HIEs may offer a promising resource for conducting population-based postmarketing surveillance of medical products with EHR data.

## Data Availability

The data that support the findings of this study are not publicly available due to privacy restrictions.

## Statements and Declarations

### Funding Acknowledgements

This project was supported by Master Agreement *75F40119D10037* from the US Food and Drug Administration (FDA). FDA coauthors reviewed the study protocol, statistical analysis plan, and the manuscript for scientific accuracy and clarity of presentation. Representatives of the FDA reviewed a draft of the manuscript for presence of confidential information and accuracy regarding statement of any FDA policy. The views expressed are those of the authors and not necessarily those of the U.S. FDA.

### Competing Interests

Dr. Khurshid and Eliel Oliveira are co-founders of Pulsar Health, a software IT company. Dr. Desai reports serving as Principal Investigator on investigator-initiated grants to the Brigham and Women’s Hospital from Novartis, Vertex, and Bayer on unrelated projects. Dr. Schneeweiss is co-principal investigator of an investigator-initiated grant to the Brigham and Women’s Hospital from Boehringer Ingelheim unrelated to the topic of this study. He is a consultant to Aetion Inc., a software manufacturer of which he owns equity. His interests were declared, reviewed, and approved by the Brigham and Women’s Hospital and MGB HealthCare System in accordance with their institutional compliance policies. All other authors have no conflicts of interest to disclose relative to this work.

## Notes

### Author Declarations

This Sentinel project is a public health surveillance activity conducted under the authority of the Food and Drug Administration and, accordingly, is not subject to Institutional Review Board oversight. Additionally, the MyHealth Access Network governance committee comprising local stakeholders and data contributors, reviewed and approved the secondary use of the data for this query.

